# Discovery and Detection of Dry Eye Disease Protein Biomarkers Using Soft Contact Lens Tear Sampling

**DOI:** 10.1101/2023.05.18.23290182

**Authors:** Robert K. Roden, Fangfang Jiang, Alyssa Nitz, Nathan Zuniga, Rebecca S. Burlett, Leena M. Patil, Holly Farnsworth, P. Christine Ackroyd, Samuel H. Payne, John C. Price, Kenneth A. Christensen

**Affiliations:** Department of Chemistry & Biochemistry, Brigham Young University, Provo, UT, 84602, USA; College of Optometry, Rocky Mountain University of Health Professions, Provo, UT, 84606, USA; Alpine Vision Center, Saratoga Springs, UT, 84045, USA; Department of Biology, Brigham Young University, Provo, UT, 84602, USA

**Keywords:** Dry Eye Disease, Tear Film, Soft Contact Lenses, Diagnostics, Mass Spectrometry, Proteomics

## Abstract

Dry eye disease (DED) affects up to half of the world’s population and causes blur, eye pain, decreased quality of life. Unfortunately, many people do not receive proper treatment due to current challenges associated with DED diagnoses. To address this issue, point-of-care (PoC) clinical tear film diagnostics are beginning to be used for rapid in-office testing. However, these diagnostics also have significant obstacles, including challenges associated with tear sampling. We previously demonstrated how soft contact lenses (SCLs) can be used to capture and concentrate tear proteins on the eye. Given the low tear volume associated with DED subjects, SCL sampling holds great potential to improve DED PoC diagnostics and facilitate prompt treatment for patients. Here we show the detection of new and established DED protein biomarkers using our SCL sampling method. Our analysis revealed associations between DED tear proteins and the immune system, ferroptosis, salivary secretion, and calcium binding proteins. We also identified correlations between clinical DED testing and proteomics, emphasizing the power and importance of translational medicine research. These experiments lay the foundation for both SCL sampled tear protein biomarker discovery as well as improved diagnostic testing to improve DED patient care.

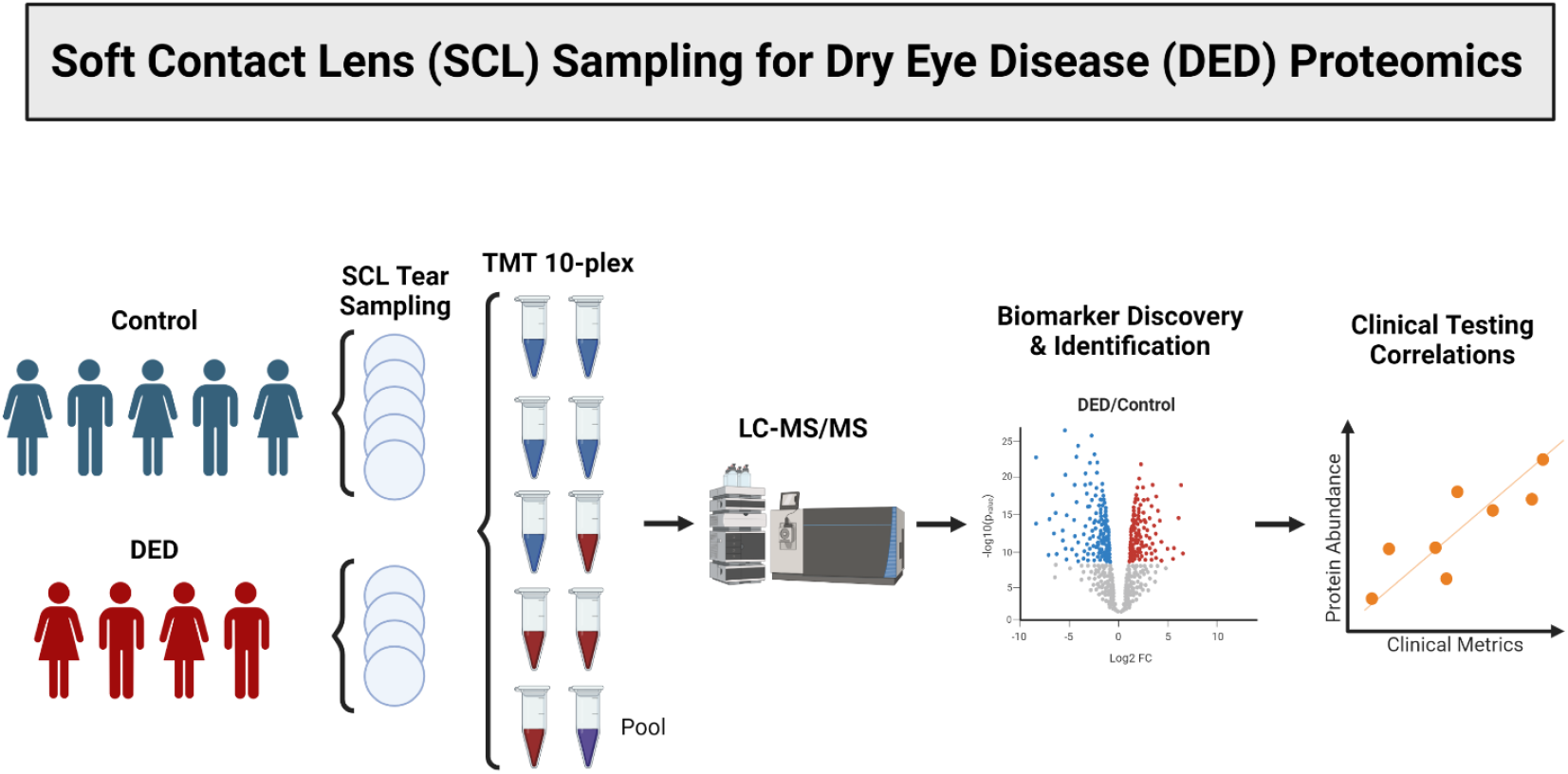

## Introduction

Dry eye disease (DED) is a common, multifactorial pathology which causes eye pain, blurred vision, and decreased quality of life [1,2]. Since accurate diagnosis is critical for optimal treatment, adequate DED diagnostics are necessary [3]. However, there is currently no gold standard test for DED and proper diagnosis is a lengthy process that requires multiple tests and significant effort [3,4]. As such, DED diagnosis is often based solely on symptoms and treatments are implemented one by one until the patient finds a satisfactory endpoint; a process which can be costly, time intensive, and frustrating for both patients and clinicians [4]. Because proper treatment and best patient outcomes depend on accurate diagnoses, improved DED diagnostic tests are needed.

Human tear film contains DED protein biomarkers and point-of-care (PoC) clinical diagnostic tests are being developed that allow for rapid and relatively non-invasive tear protein screenings [5-7]. While a variety of tear sampling methods can be used for diagnostic purposes, the most common and currently recommended tear film collection methods are Schirmer strips (SS) and microcapillary tubes (MCT) [8]. However, SS are irritating and introduce confounding proteins via reflex tearing while MCT tear sampling has is particularly challenging for DED patients due to low tear volume [9-12]. Importantly, a recent report by Krajčíková et. al states that both SS and MCT are “unsuitable” methods for DED subjects [10]. Thus, improved tear sampling methods for DED subjects are needed.

Our previous work has shown that soft contact lenses (SCLs) are a useful method for basal tear sampling which could be particularly advantageous for DED subjects [12]. SCLs are designed for optimal comfort and concentrate tear proteins on the lens, making low tear volumes in DED patients less of a problem [13]. They also allow patients to self-sample their own tears, a considerable advantage for those whose eyes are already irritated and are averse to eye touch for tear sample collection. SCL tear sampling is therefore an advantageous alternative to current tear sampling methods for DED diagnostics, though it has not yet been demonstrated.

We hypothesized that SCLs can be used for tear protein biomarker discovery in DED patients. In this pilot study, we test our hypothesis on a small group of subjects as well as analyze our data for clinical testing correlations to proteomic data. This proof-of-concept study shows that SCL sampling can be used for biomarker discovery for DED as several new and established biomarkers are identified. We also observed correlations between proteomic findings and clinical data obtained from dry eye subjects, highlighting parallels between clinical data and biochemical processes in DED. Ultimately, this study demonstrates the usefulness of SCL tear film sampling for DED patients as well as the power of translational medicine research in progressing DED diagnosis strategies and furthering predictive, preventive, and personalized medicine (3PM).

## Materials and Methods

### Human Subject Enrollment

Human subjects research was performed in accordance with the Declaration of Helsinki. Approval was granted by the Internal Review Board at Brigham Young University (IRB2022-314). Subjects were educated on the purposes, risks, and benefits of the study. Informed consent was obtained before subject enrollment and privacy rights of human subjects were observed. Enrollment was based on predetermined inclusion and exclusion criteria. Subjects who were 18 years or younger and pregnant women were excluded from the study.

We enrolled 4 males and 5 females between the ages of 18 and 87. Of these subjects, 4 were diagnosed with DED and 5 were deemed to have normal ocular health. All samples were collected and clinical testing performed at Alpine Vision Center (AVC) (Saratoga Springs, UT).

### Dry Eye Diagnosis

Determination of DED was made by an optometrist based on guidelines reported by the Ocular Surface Society (TFOS) Dry Eye Workshop (DEWS) II Diagnostic Methodology Subcommittee Report [4]. In this study, each subject answered questions about dry eye symptoms using the Ocular Surface Disease Index (OSDI) questionnaire. Next, a fluorescein dye strip (BioGlo) was wetted with BioTrue solution and gently touched to the subject’s bulbar conjunctiva. Using a Keratograph 5M, noninvasive keratograph tear breakup time (NIKBUT), meibomian gland dysfunction (MGD), superficial punctate keratitis (SPK), and tear meniscus height (TMH) were measured on each subject.

Subjects were diagnosed with DED if OSDI scores were ≥ 13 as well as a NIKBUT score < 10 seconds and/or >5 corneal or 9 conjunctival fluorescein staining spots. Subtype diagnoses was made for evaporative dry eye (EDE) vs. aqueous deficient dry eye (ADDE). EDE was diagnosed for those with any stage of meibomian gland dysfunction (MGD) and TMH > 0.2mm, while ADDE had no MGD and average TMH < 0.2mm.

### Tear sampling

Tears were sampled using etafilcon A soft contact lenses as previously described [12]. Subjects were allowed to sample their own tears under the supervision of an optometrist if they so desired.

### MS sample preparation

Samples were prepared as previously described [12], with the exception of adding 150 μL Pierce IP lysis buffer (Thermo Fisher Scientific) to the 4M guanidine solution for protein removal.

### Mass spectrometry

Online liquid chromatography–tandem mass spectrometry (LC–MS/MS) was performed using instruments, methods, and settings as previously described [12].

### Data Analysis

MS data was analyzed using Peaks software (Bioinformatics Solutions Inc.). Spectrum filter settings were set to false discovery rate (FDR) of 1% (−10logP ≥ 26.4) and quality ≥ 8.8. Protein filter settings were significance ≥ 0, fold change ≥ 1, and at least 1 unique peptide. All spectra with intensity ≤1E2 were excluded. For quantitative analysis, raw data was log_2_ transformed, “0”’s were replaced with blanks, then individual protein abundances were normalized to the average protein abundance of the pooled sample.

Shapiro-Wilk tests were used to check data normality. As most of our data was not normally distributed (Supplementary Figure 1), control and DED groups were compared by performing Mann-Whitney U-tests for all proteins in R. FDR was accounted for using the Benjamini-Hochberg method with the FDR set to 0.05. Adjusted p-values < 0.05 were considered significant. For qualitative comparisons, only proteins that were present in ≥ 3 subjects were counted. Gene ontology and functional annotation analyses were performed using the DAVID bioinformatics database with classification stringency set to medium for functional annotation clustering [14,15].

Clinical test scores were averaged between both eyes. TMH was calculated by averaging nasal, lateral, and temporal measurements. The Shapiro-wilk test was also used to test normality of clinical data. As the majority of our data was not normally distributed, statistical analysis of correlations between clinical data and protein abundance was made using Spearman correlations. Correlations were calculated using clinical scores and individual protein abundances for all subjects with Spearman values > 0.7 and < -0.7 considered significant.

The raw data is available on the Mendeley Data repository [16].

## Results

To demonstrate biomarker discovery with SCL tear sampling, we compared tear film proteins between 4 DED to 5 healthy subjects. As DED is a multifactorial pathology with many variables, clinical data was collected to better characterize the type and extent of pathology. For each subject SPK, TMH, NIKBUT, MGD, and OSDI were measured prior to tear sampling (Table 1). Based on these data, subjects who met the inclusion criteria were recruited and placed into DED and healthy control groups.

**Table 1.** Averaged Binocular Clinical DED Test Scores for All Subjects

| Clinical Test | <sup>1</sup>Subject |  |  |  |  |  |  |  |  |
| --- | --- | --- | --- | --- | --- | --- | --- | --- | --- |
|  | C1 | C2 | C3 | C4 | C5 | D1 | D2 | D3 | D4 |
| <sup>2</sup>SPK (0-4) | 0.0 | 0.0 | 0.0 | 0.0 | 0.0 | 0.0 | 1.5 | 0.0 | 0.5 |
| <sup>3</sup>TMH (mm) | 0.72 | 0.40 | 0.25 | 0.70 | 0.44 | 0.70 | 0.44 | 0.25 | 0.59 |
| <sup>4</sup>NIKBUT (1st Breakup, sec.) | 3.82 | 8.03 | 8.13 | 7.08 | 5.93 | 2.49 | 2.29 | 4.30 | 5.83 |
| <sup>5</sup>MGD (0-4) | 0.0 | 0.0 | 0.5 | 0.0 | 0.5 | 2.0 | 0.0 | 1.5 | 1.0 |
| <sup>6</sup>OSDI (0-100) | 4.17 | 2.08 | 10.42 | 0.00 | 6.25 | 35.00 | 47.92 | 45.83 | 37.50 |

<sup>1</sup> “C” designates control subjects, “D” designates DED subjects
<sup>2</sup> Superficial punctate keratitis (SPK) is scored 0 (none) to 4 (severe)
<sup>3</sup> Tear meniscus height (TMH)
<sup>4</sup> Non-invasive keratograph break up time (NIKBUT)
<sup>5</sup> Meibomian gland dysfunction (MGD) is scored 0 (none) to 4 (severe)
<sup>6</sup> Ocular Surface Disease Index (OSDI) is scored 0 (no symptoms) to 100 (extremely symptomatic)

Next, we sought to characterize DED and control proteomes to identify putative DED biomarkers using SCL tear sampling. Despite significant intersubject proteomic variation, we identified 109 DED and 99 healthy control proteins present in 3 or more subjects using LC-MS/MS (Figure 1). Of these proteins, 20 were unique to DED subjects and 10 to healthy controls (Table 2). Gene ontology analysis showed that unique DED proteins were strongly associated with the immune response, including B cell activation, phagocytosis, immunoglobulin binding, bacterial defenses, antigen binding, and complement activation (Supplementary Table 1). These proteins were also correlated with salivary secretion and calcium binding. Quantitative analysis no statistically significant differential expression between DED and control subject groups (Supplementary Table 2).

**Figure 1.**
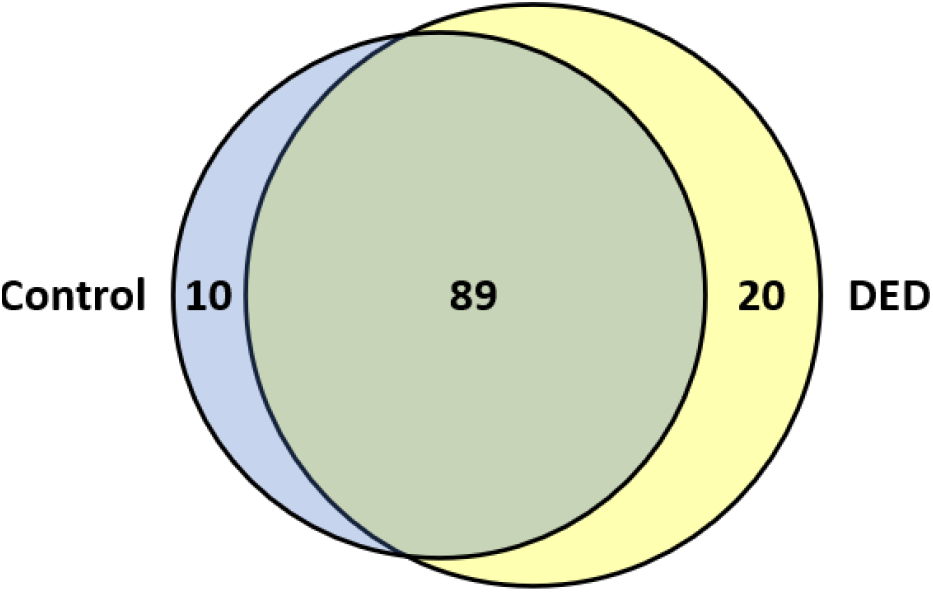
Venn Diagram of protein identifications in 3 or more subjects. Right/blue = control, Left/yellow = DED.

**Table 2.** Unique Proteins Identified in Control and DED Subjects

| Control | DED |
| --- | --- |

| Uniprot | Protein | Uniprot | Protein |
| --- | --- | --- | --- |
| P00450 | Ceruloplasmin (CERU) | P01742 | Immunoglobulin heavy variable 1-69 (HV169) |
| P01037 | Cystatin-SN (CYTN) | Q7Z794 | Keratin, type II cytoskeletal 1b (K2C1B) |
| P06396 | Gelsolin (GELS) | P0DP06 | Immunoglobulin heavy variable 4-30-4 (HVD34) |
| A0A0C4DH29 | Immunoglobulin heavy variable 1-3 (HV103) | Q9H8W5 | Tripartite motif-containing protein 45 (TRI45) |
| P0DP01 | Immunoglobulin heavy variable 1-8 (HV108) | P08582 | Melanotransferrin (TRFM) |
| P06331 | Immunoglobulin heavy variable 4-34 (HV434) | P0DTE7 | Alpha-amylase 1B (AMY1B) |
| A0A0C4DH41 | Immunoglobulin heavy variable 4-61 (HV461) | P14923 | Junction plakoglobin (PLAK) |
| P01700 | Immunoglobulin lambda variable 1-47 (LV147) | P07355 | Annexin A2 (ANXA2) |
| P12035 | Keratin, type II cytoskeletal 3 (K2C3) | Q02413 | Desmoglein-1 (DSG1) |
| P80188 | Neutrophil gelatinase-associated lipocalin (NGAL) | A0A0B4J2H0 | Immunoglobulin heavy variable 1-69D (HV69D) |
|  |  | P01619 | Immunoglobulin kappa variable 3-20 (KV320) |
|  |  | P14618 | Pyruvate kinase PKM (KPYM) |
|  |  | P49788 | Retinoic acid receptor responder protein 1 (TIG1) |
|  |  | Q92820 | Gamma-glutamyl hydrolase (GGH) |
|  |  | P05164 | Myeloperoxidase (PERM) |
|  |  | P12830 | Cadherin-1 (CADH1) |
|  |  | P02763 | Alpha-1-acid glycoprotein 1 (A1AG1) |
|  |  | P28325 | Cystatin-D (CYTD) |
|  |  | A0A0C4DH55 | Immunoglobulin kappa variable 3D-7 (KVD07) |
|  |  | P02671 | Fibrinogen alpha chain (FIBA) |

Since many DED biomarkers have been previously reported [17], we sought to identify any of them in our results. Various biomarkers were identified, including actin, cytoplasmic 1 (ACTB), serum albumin (ALB), annexin A2 (ANXA2), zinc-alpha-2-glycoprotein (AZGP1), beta-2-microglobulin (B2M), complement C3 (C3), clusterin (CLU), cystatin SN (CST1), interferon gamma-induced protein 10 (CXCL10), dermcidin (DCD), deleted in malignant brain tumors 1 protein (DMBT1), alpha-enolase (ENO1), haptoglobin (HP), lipocalin-1 (LCN1), alpha-1-acid glycoprotein 1 (ORM1), prolactin-inducible protein (PIP), S100 calcium-binding protein A8 (S100A8), secretoglobin family 2A member 1 (SCGB2A1), transcobalamin-1 (TCN1), and transferrin (TF) (Table 3). ALB, DMBT1, SCGB1D1, LACRT, PRR4, B2M, and PIP were also identified as previously reported SCL-related dry eye biomarkers. It is important to note that while we identified these biomarkers in our samples, not all reported trends in abundance concur with our findings and none were statistically significant using our predefined FDR adjustment.

**Table 3.**
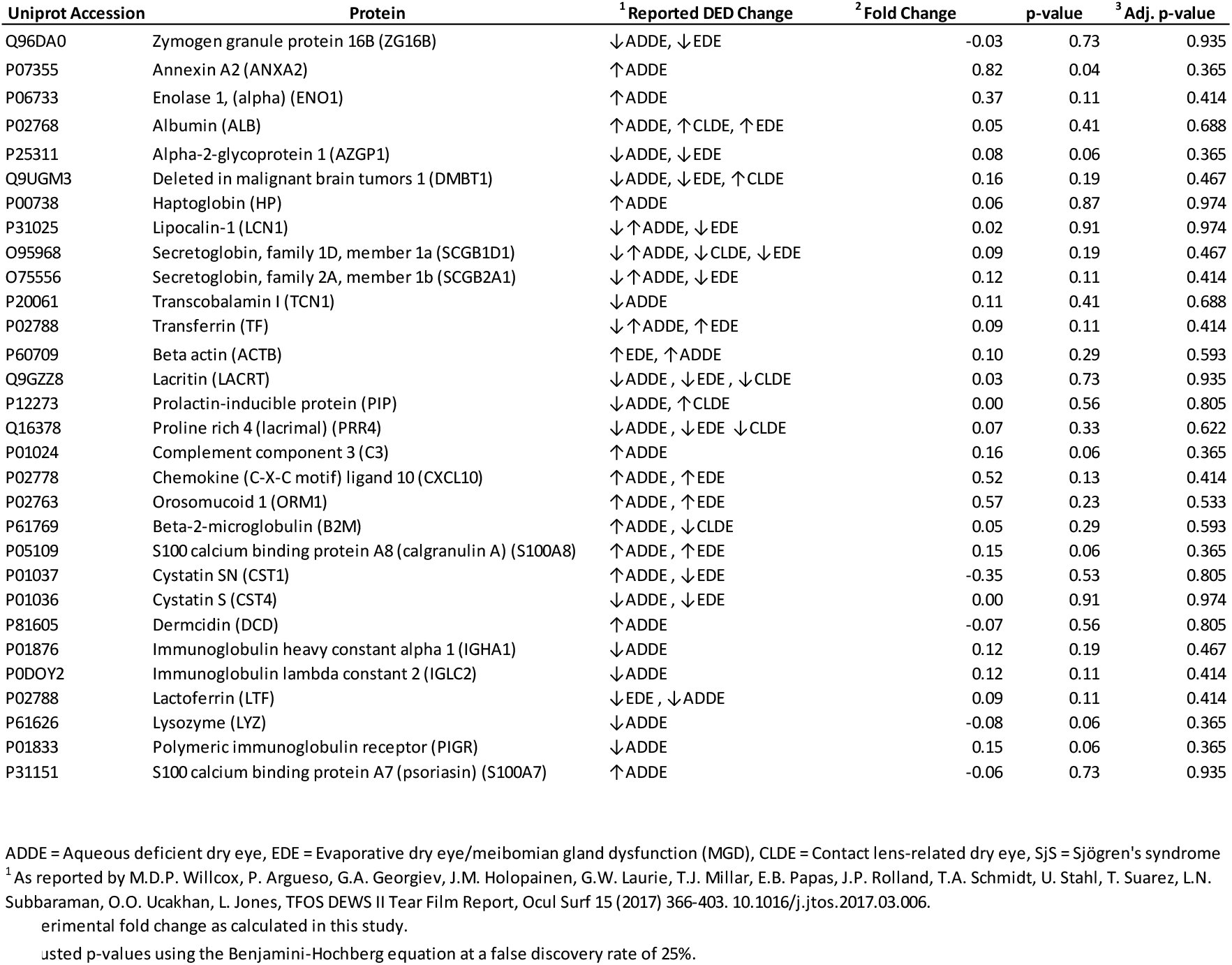
Previously Reported DED Biomarkers Identified by SCL Tear Sampling

In this study, we also sought to identify correlations between clinical and proteomic data. Interestingly, we found correlations between protein expression for most clinical tests (Table 4). Gene ontology and functional analysis further revealed correlations with secretory pathways and immune system activation (Supplementary Table 3). Significantly, we observed correlations between NIKBUT and proteins involved in ferroptosis, an iron-dependent cell death caused by excessive oxidative stress. Importantly, ferroptosis has recently been identified as a DED pathogenic mechanism [18,19]. Our data substantiates oxidative stress as a key factor in DED pathogenesis and connects proteomic data to iron-dependent mechanisms of tear film instability [20,21]. Furthermore, we identified that NIKBUT and OSDI are correlated with proteins involved with calcium binding. While calcium is known to play an important role in DED, these findings may help elucidate mechanistic details for DED associated tear film stability and ocular pain [22,23].

**Table 4.**
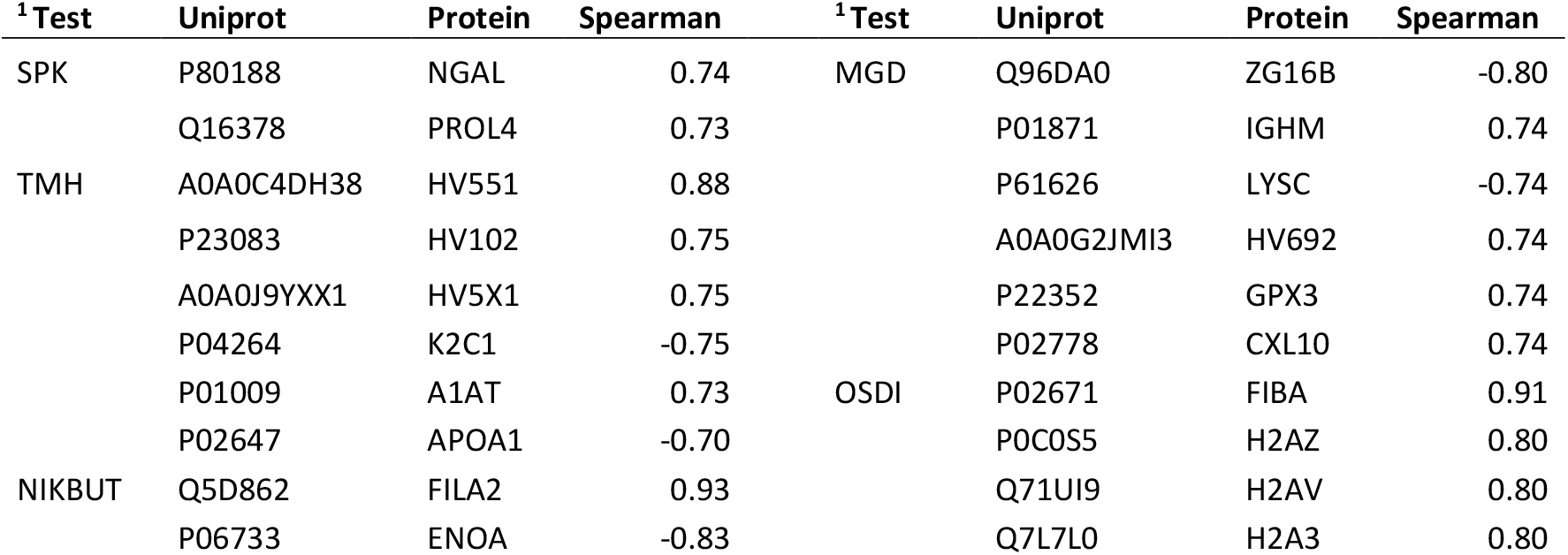
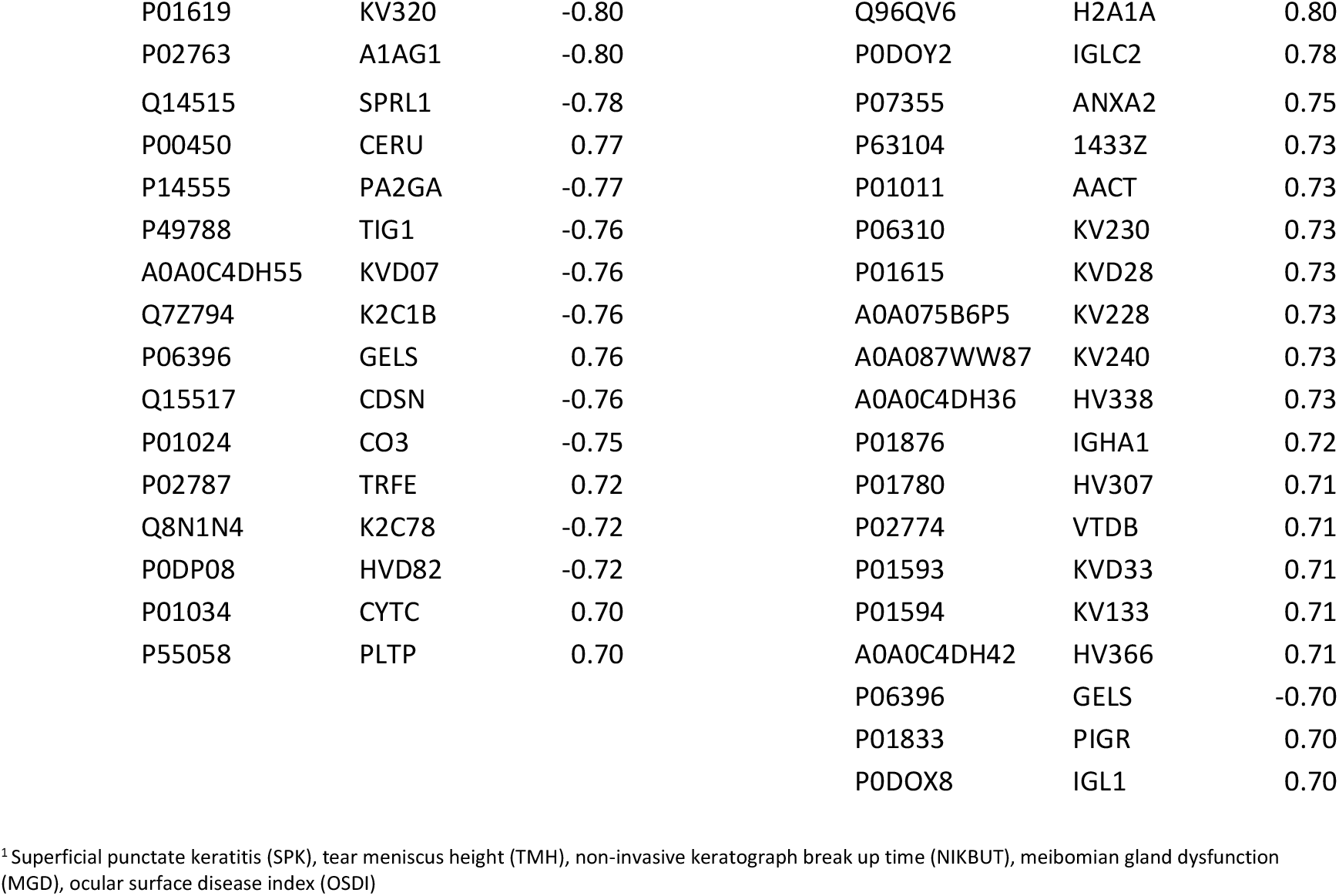
Spearman Correlations Between Proteomic and DED Clinical Testing Data

Taken together, these data demonstrate the use of SCL sampling for biomarker detection in DED subjects. They also demonstrate the use of clinical testing to understand proteomic shifts that occur with the loss of tear film homeostasis in DED.

## Discussion

Up to 50% of the world’s population is estimated to have DED [1] and over a third of DED cases in the US remain undiagnosed [24]. Thus, finding improved methods for diagnosing DED is important. To address this issue, tear protein biomarkers have recently been used to create clinical PoC diagnostic DED tests [6,25]. However, given the low tear volume inherent in DED subjects, tear sampling for these tests can be challenging. The currently recommended SS and MCT methods are irritating, and introduce confounding proteins via reflex tearing [9]. MCT sampling increases the risk of injury in DED patients given that low tear volumes may require more time to manipulate the tube on the ocular surface to collect tears [26]. In contrast, SCLs capture and concentrate tear proteins on the eye, making low tear volumes in DED patients less of a problem. They also allow patients to self-sample their own tears, a considerable advantage for those whose eyes are already irritated and don’t want other people touching their eyes. As SCL tear sampling is an advantageous alternative to current tear sampling methods for DED diagnostics, we used proteomics to quantify and identify DED biomarkers from patient samples.

In this pilot study, we identified 20 potential qualitative DED biomarkers from 4 DED and 5 control subjects using SCL sampling and LC-MS/MS. We also identified 30 previously reported quantitative DED biomarkers. We thus conclude that SCL sampling is an alternative tear sampling method for tear film protein biomarker detection. Coupled with our previous work on SCL tear sampling [12], these data also indicate the potential usefulness of SCL sampling for DED PoC diagnostics.

Since DED is a multifactorial pathology DED with up to 14 distinct subtypes, another goal of DED PoC diagnostics is specific subtype diagnosis [27]. The two main classifications of DED are evaporative dry eye (EDE) and aqueous deficient dry eye (ADDE). EDE is most commonly a result of meibomian gland dysfunction (MGD) and is commonly associated with age, allergies, and inflammation [28]. ADDE is result of decreased lacrimal gland tear production and represents only 10% of DED patients [29]. Of ADDE patients, approximately 10% of ADDE patients have Sjögren’s syndrome (SjS), an autoimmune disease affecting exocrine glands that results in dry eyes, skin, and mouth [30,31]. Mixed mechanism dry eye (MMDE) describes patients with signs of both EDE and ADDE. Because understanding individual patient DED pathophysiology determines treatment strategies and best outcomes, it is important to quickly and accurately diagnose each patient’s specific DED subtype [3].

In this study, all subjects were diagnosed with EDE based off established clinical testing procedures [4]. Interestingly, most of the biomarkers identified in our study were ADDE markers. Furthermore, we observed proteins found in our EDE subjects were associated with salivary secretion. Dry mouth (xerostomia) is strongly associated with SjS, which constitutes a major classification of ADDE. Previous studies have also shown correlations between dryness in eyes and the mouth, particularly with SjS [30,32]. Thus, proteins associated with xerostomia in these subjects may be indicative of MDDE and subclinical ADDE mechanisms, potentially including SjS [31]. While further research will be needed to understand and validate these correlations, our data shows the potential for and importance of understanding pathologic mechanisms of xerosis through tear film analysis. These data also emphasize that biochemical changes precede phenotypical manifestations observable by clinical testing. Thus, tear film biomarkers are useful for earlier diagnoses of DED and potentially other diseases as well [33]. Early-stage diagnoses using tear film diagnostics also allows for faster implementation of treatments, reduced patient suffering, and better overall outcomes [34].

Another useful insight from this study was the relationship between clinical data and proteomics. If clinical measurements are strongly correlated with proteomics, clinical tests could theoretically be used to provide biochemical insights without biospecimen sampling. Previous studies comparing clinical data to proteomics identified correlations between lactoferrin and NIKBUT, matrix metalloproteinase-9 (MMP9) and TMH, and MMP9 and bulbar conjunctival redness [35,36]. Although we did not identify MMP9 in our DED samples and observed only a weak correlation between NIKBUT and lactoferrin (Spearman = -0.57), we did observe that unique DED proteins were associated with calcium.

DED is a pathophysiologic cycle characterized by a loss of tear film homeostasis due to a variety of mechanisms, including calcium dysregulation (Figure 2). Calcium binding proteins such as psoriasin (S100A7), calgranulin A (S100A8), annexin A2 (ANXA2), and alpha-2-glycoprotein 1 (AZGP1) are established DED biomarkers [17,37] which we identified in our subject tear samples (though our observed trends in abundance did not always match previous reports). Another study by Willcox et. al also observed DED proteins associated with calcium, including alpha-2-HS-glycoprotein, annexin A2, annexin A5, calcitonin related polypeptide alpha, calreticulin, and neuropeptide Y [17]. Interestingly, all of the calcium-related proteins in their study were identified with ADDE and most were in patients with SjS. Since we identified some of the same proteins in EDE subjects, it is possible that these calcium-related DED proteins may be useful in diagnosing subclinical ADDE/MMDE.

**Figure 2.**
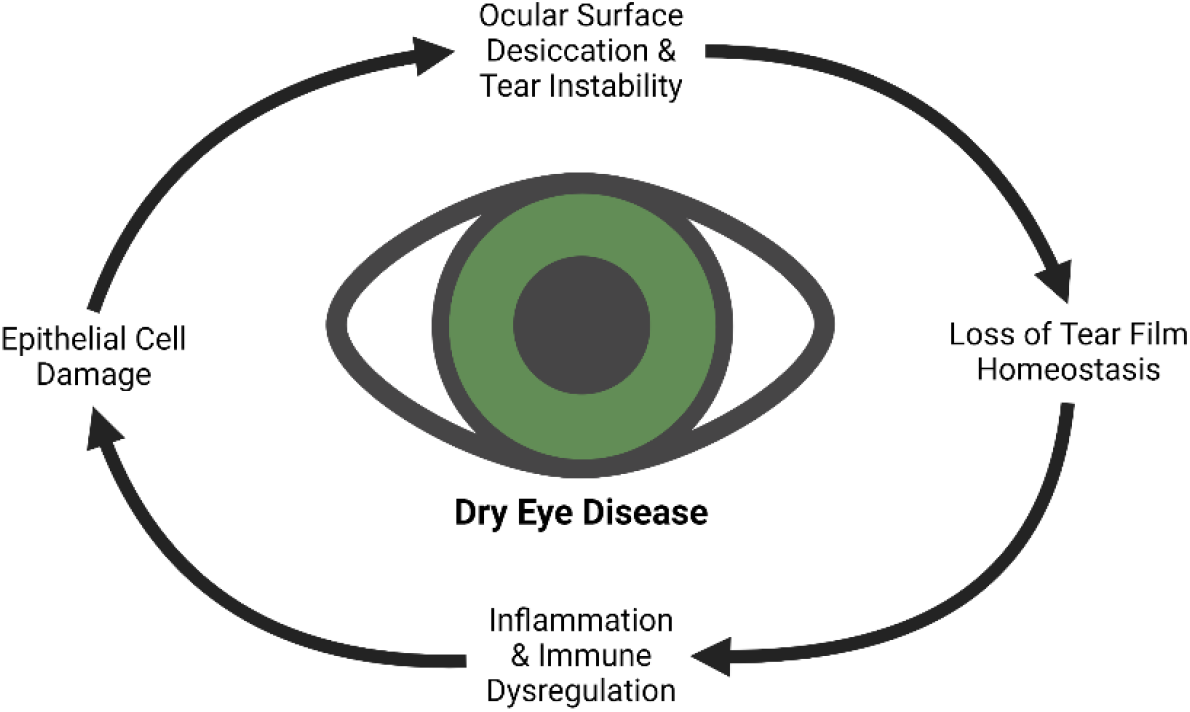
The pathophysiologic cycle of Dry Eye Disease

Interestingly, our study also showed a correlation between DED patient symptoms (including eye pain and irritation) and calcium binding proteins. Because corneal tissue is highly innervated, epithelial defects (such as SPK) associated with DED can cause severe pain. Calcium is known to directly affect corneal epithelial cells through the cytosolic iron-sulfur cluster-binding protein, CDGSH Iron Sulfur Domain 2 (CISD2). Reduced levels of CISD2 in corneal cells leads to increased calcium dysregulation via calcineurin hyperactivity, the formation of reactive oxygen species (ROS), corneal epithelial defects, and decreased wound healing [38-40]. Interestingly, DED patients who take calcineurin inhibitor cyclosporine A have shown improved corneal epithelial integrity in CISD2 knockout mice and human DED trials [40]. Furthermore, a study by Tsubota et. al found that calcium ointments were beneficial for improving DED symptoms. Though calcium treatments are not currently used to treat DED, these findings reinforce calcium’s role in DED pathophysiology and tear film stability. Our findings also show how clinical testing can provide insights into possible biochemical mechanisms of a multifactorial disease. In the future, OSDI reported symptoms may provide a way to direct treatment strategies without the need for biospecimen sampling.

Correlations were also made between NIKBUT and ferroptosis. Importantly, ferroptosis has been previously hypothesized as a key mechanism in DED pathophysiology [19]. As tears are exposed to UV light and other environmental factors, oxidative stress leads to the formation of ROS, causing the hallmark loss of tear film homeostasis and subsequent inflammation associated with DED [20]. Furthermore, the cycle of oxidative stress, ROS production, and inflammation also involves regulated cell death mechanisms, such as ferroptosis [19]. Though the mechanistic details are still being investigated, ferroptosis leads to the production of lipid peroxides [41]. Thus, it is possible that alterations of lipid molecules could affect the tear lipid layer and subsequent tear break up times as observed in our study. Importantly, CISD2 may also play an important role in connecting the iron and calcium pathways associated with DED.

Aside from proteins being related to calcium binding and ferroptosis, our analysis also revealed that proteins unique to DED were associated with immunity. This finding was expected as ocular surface damage in DED results from a loss in tear film homeostasis and dysregulated ocular immune responses [42]. Since dysregulated immunity is central to DED, steroids and non-glucocorticoid immunomodulator drugs (such as cyclosporine A) are standard and effective treatments for many DED patients [3,42]. However, since DED is a multifactorial pathology, these medications are not effective for all DED subtypes. Therefore, the biomarkers identified in this study could be useful for directing targeted immunomodulatory treatment strategies in conjunction with 3PM diagnostics [43].

While we were able to identify new and established DED biomarkers, it is worth mentioning that some previously reported biomarkers were not confirmed or did not align with our study [17]. However, it is important to note that unvalidated biomarker discrepancies for DED are common in the literature and approximately half of all DED markers have not been validated by large scale studies [7,35,37,44]. Other possible explanations for discrepancies can be associated with limitations of the study, including, 1. Our study group was small and individual tear proteomes can vary substantially, even between eyes in the same subject [35] 2. All of our subjects were of the same race and relative location, 3. We did not have age-matched controls (in large part due to the strong correlation between increasing age and DED progression), and 4. Differences in sampling method are known to affect relative protein abundance [11,12]. These limitations emphasize the need for larger studies with increased statistical power to better understand protein changes in DED patients.

Although larger studies will be needed to validate these as true DED biomarkers, this study confirms that SCLs can indeed be used for biomarker discovery. Based on our previous research, we know SCL sampling is particularly useful for DED patient tear sampling for research and clinical diagnostics. Additionally, the correlations we identified between clinical data and proteomics highlight the central role of the immune system in DED and associated xero-pathologies. Clinical testing may also provide early insights into biochemical mechanisms of DED without the need for invasive testing. Such technology would allow for more specific and targeted DED treatments, consistent with the goals and benefits of 3PM.

## Data Availability

All data produced are available online at Mendeley Data

https://data.mendeley.com/datasets/jnxndrvgw2/draft?a=37cf3cbd-0544-4f09-9f44-f0f00320fc0a

## Statements and Declarations

## Acknowledgements

We would like to thank Dr. Steven Weaver and Dr. Carlan Reese at Alpine Vision Center.

## Ethics approval

Approval for research involving human subjects was granted by the Internal Review Board at Brigham Young University (IRB2022-166).

