## Supplementary Materials (Figures) for "Discovery and Detection of Dry Eye Disease Protein Biomarkers Using Soft Contact Lens Tear Sampling"

### Supplementary Information

**Supplementary Table 1**
*Gene Ontology and Functional Analyses of DED and Healthy Control Proteins*

**(Excel File)**

**Supplementary Table 2**
*Quantitative Protein Changes Between Dry Eye and Control Subjects*

**(Excel File)**

**Supplementary Table 3**
*Gene Ontology and Functional Analyses of Proteins with Significant DED/Clinical Testing Correlations*

**(Excel File)**

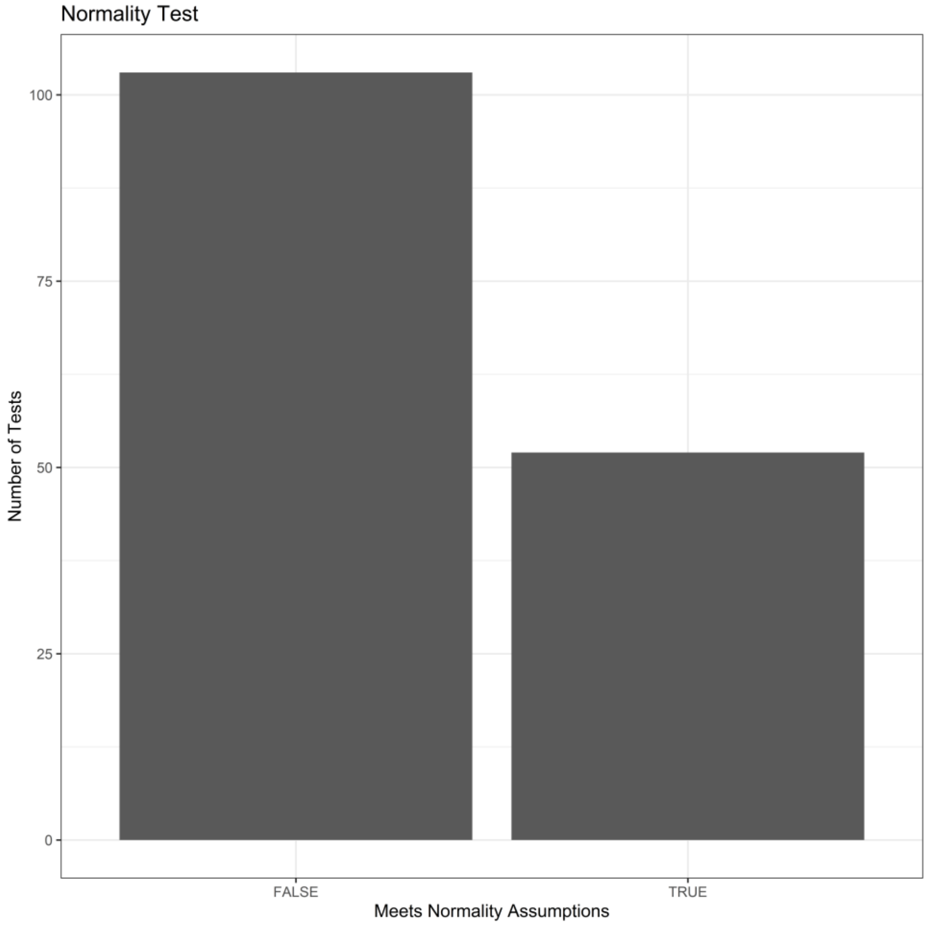


**Supplementary Figure 1.** Shapiro-Wilk normality test for proteins identified by LC-MS/MS
